# Context matters: Contrasting behavioral and residential risk factors for Lyme disease between two high-incidence regions in the Northeastern and Midwestern U.S

**DOI:** 10.1101/2020.01.31.20019810

**Authors:** Gebbiena M. Bron, Maria del P. Fernandez, Scott R. Larson, Adam Maus, Dave Gustafson, Jean I. Tsao, Maria A. Diuk-Wasser, Lyric C. Bartholomay, Susan M. Paskewitz

## Abstract

The dynamics of zoonotic vector-borne diseases are determined by a complex set of parameters including human behavior that may vary with socio-ecological contexts. Lyme disease is the most common vector-borne disease in the United States and the Northeast and upper Midwest are the regions most affected - two areas with differing levels of urbanization and sociocultural settings. The probability of being diagnosed with Lyme disease is related to the risk of encounters with an infected blacklegged tick, which reflects both the environmental tick hazard and human behaviors. Herein, we compare behavioral and peridomestic risk factors associated with human-tick encounters between high-incidence states in the Northeast (New York and New Jersey) and Midwest (Wisconsin) of the United States. We used a smartphone application, The Tick App, as a novel survey tool, during spring and summer of 2018. Adaptive human behavior was identified in the relationship between outdoor activities and the use of preventive methods. More frequent recreational outdoor activities and gardening (a peridomestic activity) were associated with an increased likelihood of using personal protective measures. Weekly participation in non-seasonal recreational and peridomestic outdoor activities in spring and summer was associated with an increased likelihood of finding a tick in the fall or winter. Most outdoor activities were more frequently reported by participants from the Midwest than the Northeast. Participants in the Northeast reported less use of personal protective measures, but they reported more interventions to reduce the presence of peridomestic deer and ticks (i.e. pesticide applications on their property) than participants in the Midwest. Participants from the Midwest were more likely to kill rodents on their property. Context mattered, and our study illustrates the need for the assessment of personal behavior and tick exposure in these two Lyme disease-endemic regions to aid in targeted public health messaging to reduce tick-borne diseases.

**Highlights:**

- Use of personal tick prevention was associated with more frequent outdoor activity
- Personal protective measure use was higher in the Midwest than Northeast
- Interventions reducing peridomestic deer and ticks more common in the Northeast

## Introduction

Lyme disease is the most common vector-borne disease in the United States and represents over 80% of reported tick-borne disease cases (Rosenberg et al., 2018). The causative agent, *Borrelia burgdorferi* sensu stricto (Burgdorfer et al., 1982), is transmitted by two tick species, *Ixodes scapularis* (Say) and *I. pacificus* (Cooley and Kohls) in the United States. The geographic range of *I. scapularis* has been expanding (Eisen et al., 2016) with predicted establishment of the enzootic transmission of the pathogen and Lyme disease cases approximately three to five years later (Ogden et al., 2013). Humans are incidental hosts of *B. burgdorferi*, and infection occurs after exposure to an infectious tick (typically a nymph which are small and active in late spring and early summer), normally encountered outdoors, in proximity to wooded environments and during recreational, work-related or peridomestic activities (Porter et al., 2019; Stafford and Magnarelli, 1993).

Spatial patterns of Lyme disease risk (i.e. the likelihood of acquiring the disease) in the United States are geographically clustered and dynamic, with high incidence states located in the Northeast and Midwest (Kugeler et al., 2015). These high-risk clusters correlate with increased hazard (‘the source of harm’), measured as the density of host-seeking *I. scapularis* nymphs (Diuk-Wasser et al., 2006; Eisen et al., 2016). However, while tick density is a predictor of disease risk at a national scale, this relationship varies in strength at the county level (Pepin et al., 2012). These variations have been, in part, attributed to potential differences in human behaviors that play a critical role in determining human exposure to the hazard, or in mitigating its negative effects by engaging in risk reduction practices (Eisen and Eisen, 2016). Studies assessing human behavioral risk factors have mostly been local (among others: Connally et al., 2009; Orloski et al., 1998; Smith et al., 2001; Vázquez et al., 2008), precluding regional comparisons. To overcome this limitation, we developed a smartphone application, The Tick App, to conduct standardized surveys on human exposure and behavior across regions in a cost-efficient manner (Fernandez et al., 2019). Study participants self-administered a survey on the behavioral and environmental risk factors of contracting tick-borne diseases, thus providing new insights into drivers of human risk that are superimposed on or interact with the hazard.

Without a human vaccine or community-based interventions, Lyme disease prevention relies on personal protective behaviors during outdoor activities and interventions in peridomestic settings targeting the enzootic cycle to reduce ticks or pathogen transmission (Schiffman et al., 2016). Personal protective behaviors include preventive practices that reduce tick exposure (e.g., avoiding risky habitats and using repellents), and practices that reduce the risk of Lyme disease after tick exposure (e.g., checking for ticks and showering after being outdoors) (Connally et al., 2009; Eisen and Dolan, 2016; Gould et al., 2008; Jones et al., 2018). Peridomestic interventions aiming at reducing the tick hazard include the applying area-wide acaricides, treating wild animal (rodents) host to kill attached ticks, and performing landscape modifications to reduce deer visitations (e.g. by using tall fencing and limit resource provisioning) and limit rodent habitat (Connally et al., 2009; Hinckley et al., 2016; Orloski et al., 1998).

According to the health belief model (Rosenstock, 1974), actions to prevent Lyme disease would be taken when individuals are knowledgeable, perceive the risk and its severity, understand the benefits of and have the self-efficacy to carry out interventions, and receive external cues to act. Thus, the ecological (e.g. landscape structure, tick hazard) and social context (e.g. experiences of friends and family, public health messaging) can influence the uptake of risk reduction practices, as humans adapt their behavior in response to tick densities and perceived risk (Berry et al., 2018). For example, implementation of personal protective behaviors by park visitors varied across an urban-to-rural gradient in Missouri (Bayles et al., 2013). Visitors to rural and exurban parks (beyond the urban fringe) were more likely to implement tick checks and utilize tick repellent compared to visitors to suburban parks while the latter were more likely to avoid tick habitat; this variation was linked to differences in the intended recreational activity of park visitors (Bayles et al., 2013). Regional differences in the socio-ecological context, such as the higher urbanization levels, population densities and median household income in the Northeast compared to the Midwest (U.S. Census Bureau 2013) may simultaneously affect residential (i.e., peridomestic features associated with tick hazard) and behavioral risk factors for Lyme disease (i.e. activity patterns, the use of personal protective behaviors and the implementation of peridomestic interventions).

In this study, we used information derived from self-administered surveys completed by users of The Tick App to compare the use of personal protection measures, the frequency of different types of outdoor activities, the implementation of peridomestic interventions, and residential risk factors associated with peridomestic tick hazard between two high-incidence regions for Lyme disease in the United States: Wisconsin (Midwest), and New Jersey and southern New York (Northeast). Additionally, to better understand the drivers behind the use of personal protective behaviors, we evaluated the association between the adoption of these behaviors and the frequency of outdoor activities, considering both recreational and peridomestic exposure scenarios and adjusting for regional differences, demographic factors, and previous Lyme disease diagnoses (e.g. previous personal experience). In the peridomestic exposure scenario, we also assessed whether hazard reduction practices (i.e., the implementation of peridomestic interventions) affected the use of personal protective behaviors.

## Material and methods

### The Tick App project

The Tick App was developed by the Midwest and Northeast Centers of Excellence for Vector-borne Diseases in collaboration with the University of Wisconsin – Madison Center for Health Enhancement System Studies (CHESS) to serve as a research tool to better understand human behaviors affecting tick exposure and engage the general public in active tick prevention across the United States (Fernandez et al., 2019). As a research tool, it includes epidemiological surveys and allows for real-time assessment of people’s locations and activities (Fernandez et al., 2019). The Tick App included a one-time enrollment survey which was designed to take less than 10 min to fill out, and aimed to retrospectively document the users’ demographic data, past experiences with ticks and tick-borne diseases, and residential and behavioral risk factors (Fernandez et al., 2019). This app was freely available through Google Play and the App Store. Participants also had the options of enrolling and completing the survey online through The Tick App website (www.thetickapp.org), completing the survey in person, or downloading the survey from the website and mailing. The enrollment survey was accessed by users upon completion of the consent form in The Tick App, online (UW-Madison Qualtrics Survey Hosting Service, Qualtrics XM, Provo, UT) or on paper. This work was conducted in accordance with Institutional Review Board approved protocols (2018-84 University of Wisconsin – Madison and AAA3750-M00Y01 Columbia University) and HIPAA regulations.

### Participants

Participants in the Northeast and upper Midwest of the United States were recruited using passive recruitment efforts through social media (Facebook, Twitter), by posting flyers and posters in public spaces, and through newspaper, television and radio interviews. Efforts were focused particularly in Wisconsin and southern New York. Active recruitment was also conducted during house visits coupled with ongoing field research involving tick sampling in yards at selected study sites (Eau Claire, WI and Staten Island, NY). During these visits, the researchers explained the objective of the app and invited residents to participate as users. Any adult over 18 years old residing in the selected regions was eligible to participate in the study. The app was available in Google Play on May 8^th^ and in App Store on May 9^th^, 2018, respectively. The Tick App promotional activities were launched during Memorial Day weekend (May 25^th^ to 28^th^), and recruitment of participants continued throughout the duration of the spring and summer of 2018 (Fernandez et al 2019).

### Survey

The enrollment survey consisted of five sections: 1) User profile, 2) Tick exposure, 3) Outdoor activities, 4) Property features, and 5) Pets. The sections captured the following information: 1) The user profile queried demographic information including gender, age, and address (Supp. Text A, questions 1-4). 2) The tick exposure section assessed the use of personal protective behaviors (wear permethrin-treated clothing, shower or bathe to remove ticks, adjust clothing or wear light-colored clothing, use tick repellent, check for ticks, or other to-be-specified measures), tick exposure during the previous fall and winter, and previous diagnosis with Lyme disease or another tick-borne disease by a physician (Supp. Text A, questions 5-7). 3) This section captured occupational, peridomestic, and recreational outdoor activities. Questions were designed to identify whether people worked or volunteered outdoors and for what duration, and to document the frequency of recreational activities (hunting, fishing, bird watching, hiking/walking/biking/running on nature trails, camping, and visiting the beach of an ocean, lake or river) and peridomestic outdoor activities (mowing the lawn and gardening) (Supp. Text A, question 8 and 9). The frequency of recreational and peridomestic outdoor activity was reported as never, about once in the summer, at least once a month, or at least once a week. 4) This section captured property features: including residential risk factors for Lyme disease, ranging from property characteristics to the presence of deer, as well as interventions to modify deer or rodent activity, and interventions to reduce tick hazard by host-targeted or environmental pesticide applications (Supp. Text A, questions 10-15). 5) The final section pets, identified the number of cats and dogs and use of pet-related tick protection (Supp. Text A, questions 16-25).

Responses were formatted as binary (Yes or No), five-point frequency scales, select the applicable answer (i.e. drop-down menu), check all that apply, or free answer forms when appropriate (name, address, text detail for an “other” response). A few exceptions were made to reduce the complexity of choice tables and to make frequencies appropriate to the question; most notably, a 4-point scale (never, about once a summer, at least once a month, at least once a week) was used to assess outdoor activity options. This study followed the recommended guidelines for reporting results of internet e-surveys (Eysenbach, 2004), the checklist for reporting results of internet e-surveys (CHERRIES) is available in Supp. Text B.

### Data analysis

We included enrollment surveys submitted between May 8^th^ and ^rd^, 2018 in our analyses (Fernandez et al., 2019). This period captured three major holiday weekends in the United States, included our peak enrollment period and high risk season for acquiring pathogens that cause Lyme disease in the US. We compared southern New York, New Jersey, and Wisconsin as the majority of Tick App participants lived in these areas (Fernandez et al., 2019): within the Midwest, 82.3% of participants lived in Wisconsin, whereas within the Northeast 76.9% of participants lived in southern New York and New Jersey, consistent with the area of influence of our study and recruitment efforts. When comparing both areas, we kept the regional reference (Midwest and Northeast) for simplicity throughout the text but we do not intend to extrapolate to the entire region. Survey responses that did not include state or residence, use of personal protective measures, outdoor activity, and property related questions were removed. In addition, those who reported an age between 7 and 18 years old were removed. A reported age of 6 or younger was assumed to be incorrect; these surveys were retained but we manually replaced the age with ‘not answered’. We checked for congruence between state and reported zipcodes (and address, if needed) to confirm that the selected state was the actual state of residence. Personal identifiable information was removed and working files for data analysis were created to preserve confidentiality of the dataset. Peridomestic host-targeted interventions were re-coded to binary variables; a ‘yes’ was assigned if any intervention method was used and ‘no’ if all responses to the intervention methods were not used (Supp. Text A question 13 and 15).

Data analysis was completed in R Statistical Computing Software (R Core Team, 2018). Responses were summarized, and a comparison between the regions of interest was made using Pearson’s Chi-squared tests without continuity correction after missing values were removed. Odds ratios and 95% confidence intervals were estimated for the likelihood of having found a tick in the prior fall or winter for each outdoor activity frequency with ‘never’ as the reference level using *oddsratio*.*wald* from package *Epitools* (Aragon, 2017).

### Modeling personal protective behaviors

Multinomial logistic models were used to assess the likelihood of reporting each of the four most commonly reported personal protective behaviors against not using personal protective behaviors (‘None’) as the reference level, depending on the frequency of a given recreational activity, the region (Midwest and Northeast), previous self-reported Lyme diagnosis (Yes or No), gender, age category, and the interaction between region and activity. The ordinal indices for frequency of each outdoor activity were transformed into numeric variables (never = 0, at least once a week = 3) for use in the multinomial logistic models allowing us to assess direct proportional association between the use of prevention strategies and the frequency of activities. We conducted a separate model for each of the recreational activities.

For those participants living in houses with yards, we also assessed the likelihood of using personal protective behaviors depending on the frequency of peridomestic outdoor activities, peridomestic tick interventions (insecticide use, interventions reducing deer activity and interventions reducing rodent activity), and accounting for frequent recreational outdoor activities with the outdoor index (Fernandez et al. 2019), the region (Midwest and Northeast), self-reported Lyme diagnosis (Yes or No), gender, and age category. Participants who worked and/or volunteered outdoors also reported frequent (monthly and weekly) outdoor activities (Fernandez et al., 2019); because these parameters were highly correlated working / volunteering outdoors was not included in the models. Tick exposure in fall and winter was not included as a co-variate as this was strongly correlated with participation in outdoor activities (see results).

For these analyses, we used the function *multinom* from R package *nnet* (Venables & Ripley, 2002). Multi-model selection was used to assess all possible model combinations and account for model selection uncertainty, by using the function *dredge* from package *MuMIn* (Barton, 2018). The odds ratios for each explanatory variable were calculated by averaging model estimates weighted by AICc using function *model*.*avg* from package *MuMIn*.

## Results

### Participants

A total of 1,093 enrollment surveys were included in the analysis, including 396 from New York and New Jersey and 697 from Wisconsin (Table 1). The Tick App was most commonly used to complete the survey (n=999). The gender distribution was similar in the Midwest and Northeast, and male and females were nearly equally represented (Table 1). The age distribution was bimodal (Fernandez et al. 2019), and the representation in age categories varied between the two study populations: 35-44 year-olds were most represented in the Northeast (28%) whereas older than 55-64 year-olds were most represented in Wisconsin (24.3%, Table 1). Although in both regions most of the participants lived in a house with yard (n=899), the proportion of participants living in an apartment was higher in the Northeast than in the Midwest (Table 1). By contrast, participants from the Midwest were more likely to have an outdoor occupation and own at least one dog (Table 1). For those who worked outdoors, the time committed to outdoor work was similar between the two regions (Table 1).

**Table 1:**
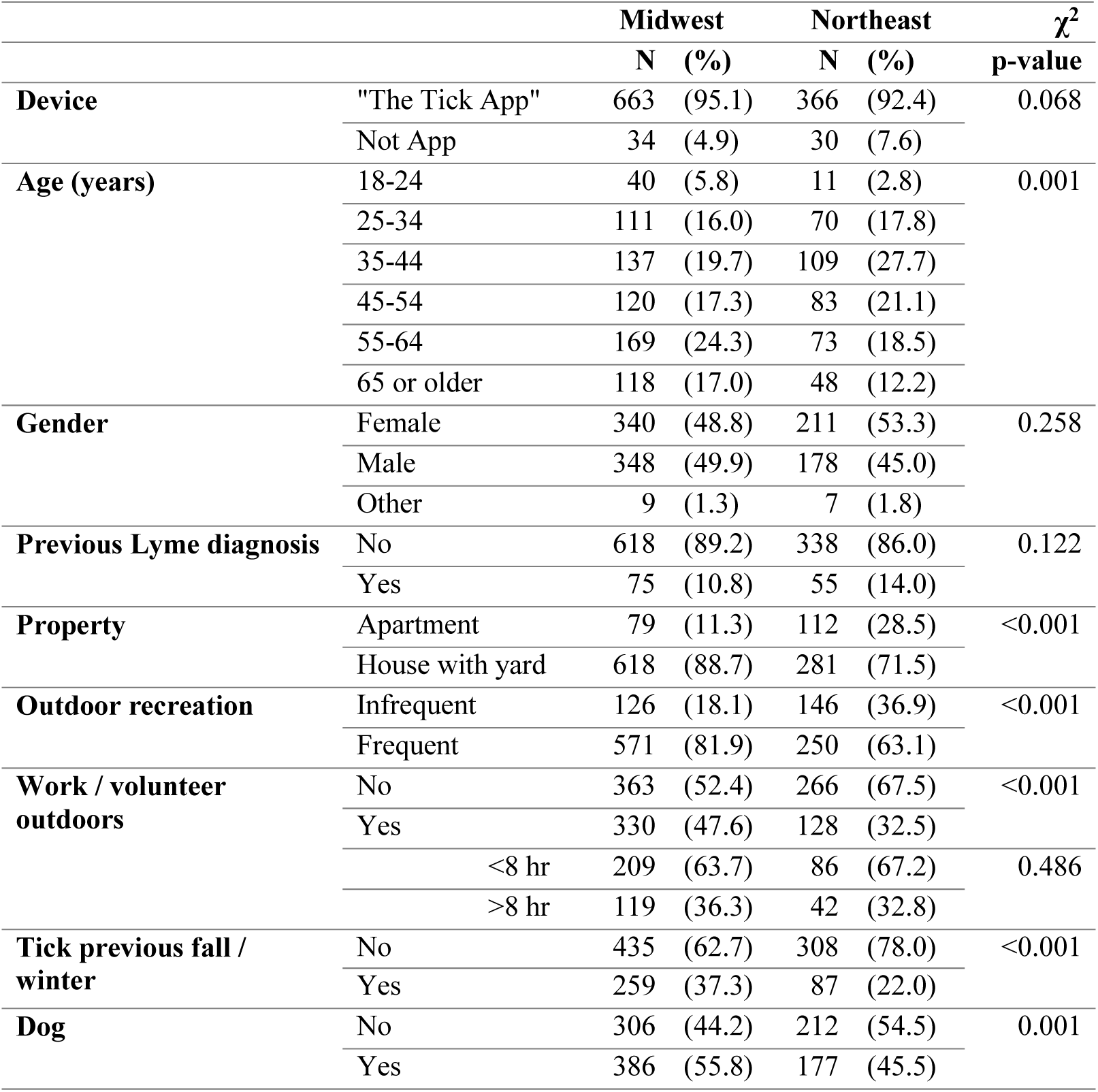
Summary of study participants. The Midwest represents participants from Wisconsin and the Northeast represents participants from New Jersey and New York.

The proportion of participants who reported a previous diagnosis of tick-borne disease was 13.6% (148 of 1089) and this was similar between participants from the Midwest (12.2% of 696) and the Northeast (16.0% of 393) (χ^2^ test, df=1, χ^2^=3.119, p=0.08). Lyme disease was most frequently reported (12.0% of 1,086, Table 1) and both babesiosis (1.8% of 1,052 respondents) and anaplasmosis (0.95% of 1,050) were rarely reported. Other diseases reported (2.0% of 1,026) included ehrlichiosis (n=6), Rocky Mountain spotted fever (n=2) and Colorado tick fever (n=1). About a third of participants (31.7%, n=1,089) found a tick on themselves during the previous fall or winter and this was significantly higher in the Midwest than the Northeast (37.3% versus 21.8%, respectively; χ^2^ test, df=1, χ^2^=27.164, p<0.001).

### Personal protective measures

The four most commonly reported personal protective behaviors were ‘Check myself for ticks’ (the most common behavior), ‘Tick repellent (e.g. DEET, picaridin)’, ‘Wear protective clothing (e.g. light colored, long-sleeved, tucking pants in socks, boots, not including permethrin-treated clothing)’, and ‘Shower or bathe to remove ticks’ (Figure 1). Midwest respondents were more likely to report that they checked for ticks, used repellent, or showered and bathed (Figure 1). Less than 15% of the study participants reported use of permethrin-treated clothing, and 4.1% reported other strategies to protect against tick bites including the use of essential oils (n=10) or staying away from grass, trees, and woods (n=10).

**Figure 1:**
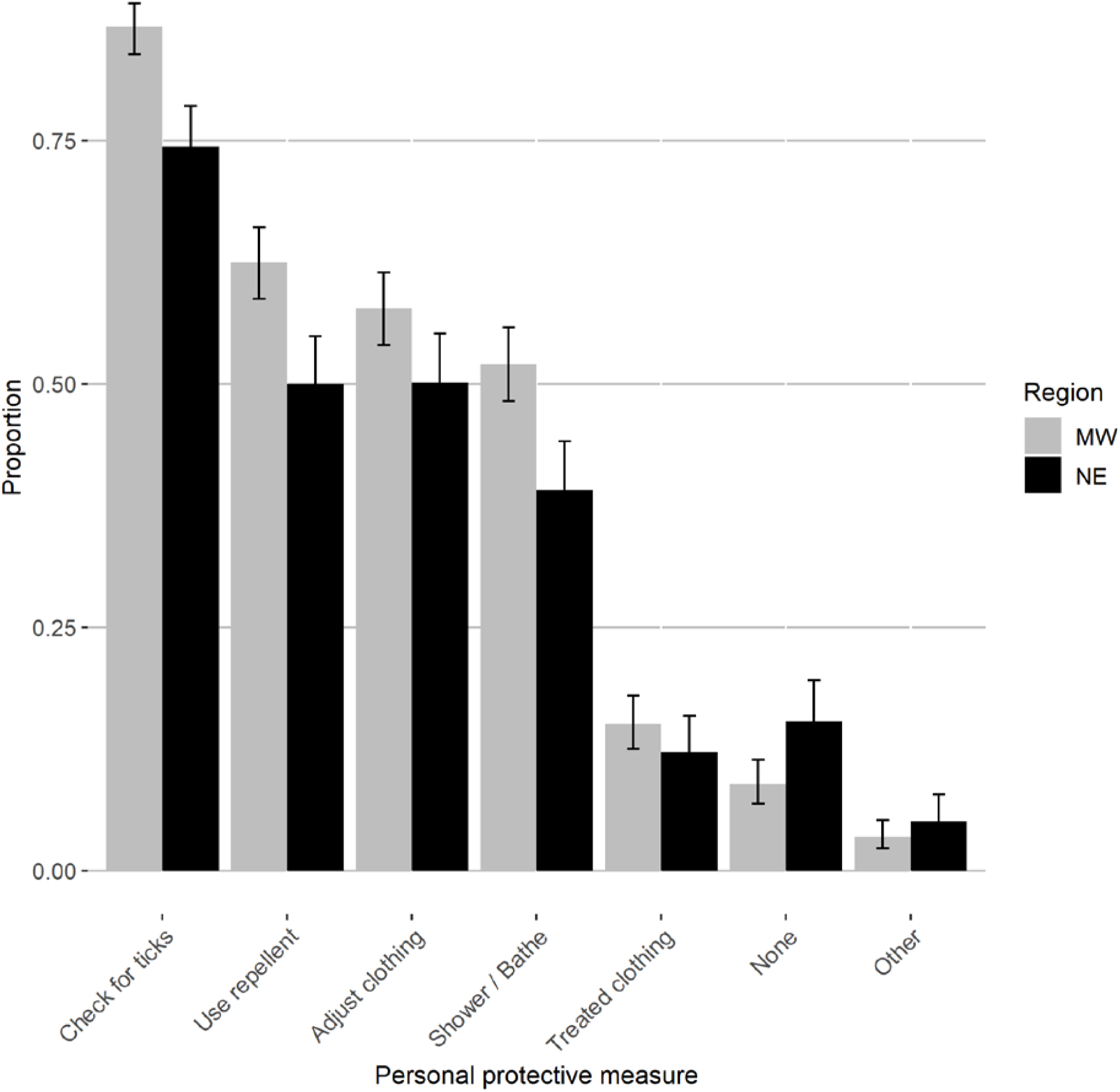
Use of personal protective measures vary between the Midwest and the Northeast. The proportion of participants who used personal protective measures the previous spring and summer. Personal protective measures included: Check one-self for ticks, use of tick repellent use (e.g. DEET, picaridin), wear protective clothing (e.g. light colored, long-sleeved, tucking pants in socks, boots), shower or bathe to remove ticks, permethrin-treated clothing, no personal protective measures and other methods. Error bars represent 95% confidence intervals.

### Recreational outdoor activities

Nearly all participants engaged in at least one of the recreational outdoor activities during the spring and summer (98.4%); only 16 of 1,060 participants responded “never” to all six recreational activities. Participation in recreational outdoor activities was greater in participants from the Midwest than the Northeast, except for visiting the beach on a lake, river, or ocean (Table 2). The odds of self-reported tick encounter during the previous fall or winter were higher if any of the recreational outdoor activities were reported to be done at least monthly or weekly in the spring and summer compared to never or once (Table 3).

**Table 2.** Midwesterners do more frequent recreational and peridomestic activity than Northeasterners. The number of respondents (n) and percentage (%) in each frequency per recreational and peridomestic outdoor activity. A comparison between the Midwest and Northeast was made for the participants in each activity frequency, the p-value of the Chi-squared test (χ^2^) is included.

| | | Midwest | | Northeast | | $\chi^2$ |
| --- | --- | --- | --- | --- | --- | --- |
| Activity | Frequency | n | (%) | n | (%) | p-value |
| *Recreational* |  |  |  |  |  |  |
| Bird watching | Never | 363 | (53.0) | 254 | (65.5) | <0.001 |
|  | About once in the summer | 82 | (12.0) | 51 | (13.1) |  |
|  | At least once a month | 91 | (13.3) | 40 | (10.3) |  |
|  | At least once a week | 149 | (21.8) | 43 | (11.1) |  |
| Camping | Never | 262 | (38.2) | 233 | (59.9) | <0.001 |
|  | About once in the summer | 217 | (31.7) | 115 | (29.6) |  |
|  | At least once a month | 176 | (25.7) | 34 | (8.7) |  |
|  | At least once a week | 30 | (4.4) | 7 | (1.8) |  |
| Fishing | Never | 296 | (43.0) | 299 | (76.7) | <0.001 |
|  | About once in the summer | 149 | (21.6) | 47 | (12.1) |  |
|  | At least once a month | 173 | (25.1) | 30 | (7.7) |  |
|  | At least once a week | 71 | (10.3) | 14 | (3.6) |  |
| Hiking | Never | 37 | (5.4) | 45 | (11.4) | <0.001 |
|  | About once in the summer | 62 | (9.0) | 71 | (18.0) |  |
|  | At least once a month | 230 | (33.4) | 119 | (30.1) |  |
|  | At least once a week | 360 | (52.2) | 160 | (40.5) |  |
| Hunting | Never | 539 | (79.1) | 373 | (95.4) | <0.001 |
|  | About once in the summer | 43 | (6.3) | 5 | (1.3) |  |
|  | At least once a month | 63 | (9.3) | 8 | (2.0) |  |
|  | At least once a week | 36 | (5.3) | 5 | (1.3) |  |
| Visiting the beach | Never | 49 | (7.1) | 24 | (6.2) | 0.911 |
|  | About once in the summer | 156 | (22.7) | 90 | (23.1) |  |
|  | At least once a month | 293 | (42.6) | 163 | (41.9) |  |
|  | At least once a week | 189 | (27.5) | 112 | (28.8) |  |

| *Peridomestic* |  |  |  |  |  |  |
| --- | --- | --- | --- | --- | --- | --- |
| **Gardening** | Never | 72 | (11.8) | 60 | (21.5) | <0.001 |
|  | About once in the summer | 38 | (6.2) | 20 | (7.2) |  |
|  | At least once a month | 98 | (16.1) | 52 | (18.6) |  |
|  | At least once a week | 401 | (65.8) | 147 | (52.7) |  |
| **Mowing the lawn** | Never | 120 | (19.6) | 147 | (53.3) | <0.001 |
|  | About once in the summer | 23 | (3.8) | 11 | (4.0) |  |
|  | At least once a month | 99 | (16.2) | 45 | (16.3) |  |
|  | At least once a week | 370 | (60.5) | 73 | (26.4) |  |

**Table 3.**
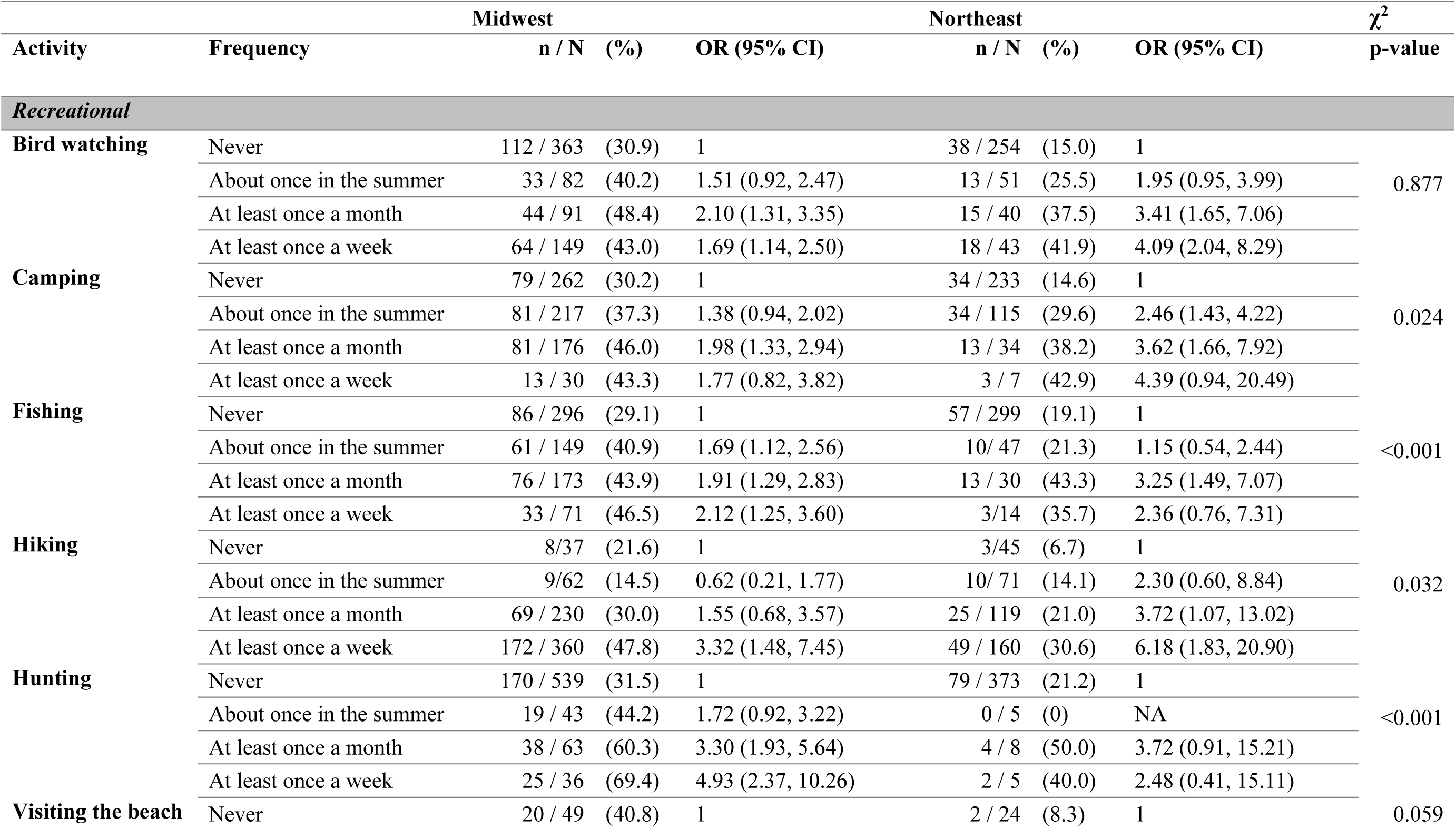
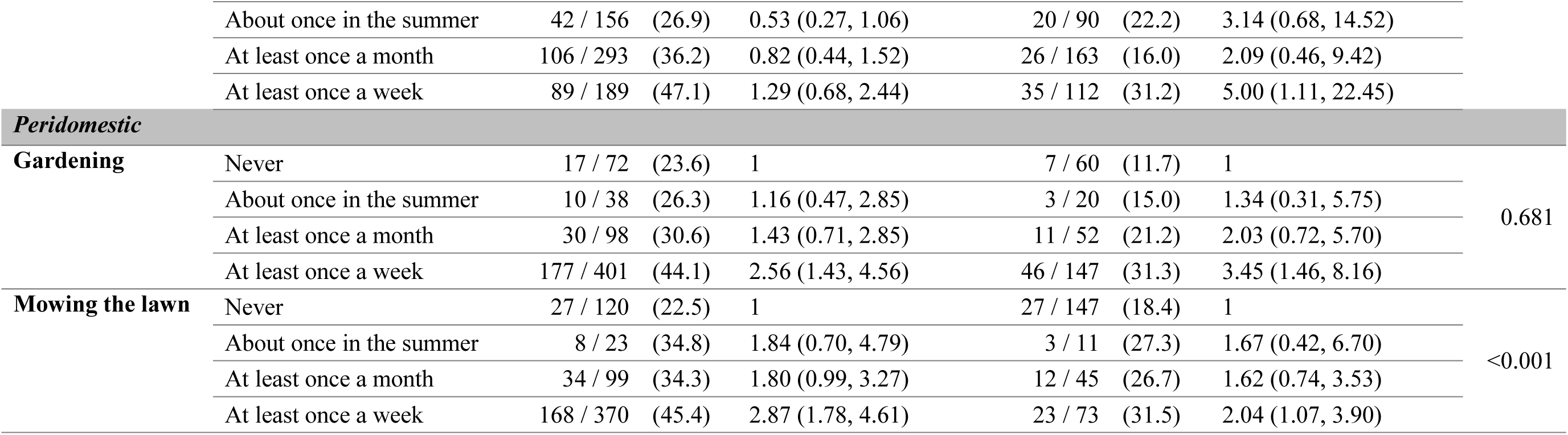
Participants who reported more frequent outdoor activity in spring and summer were more likely to have found a tick in the previous fall and winter. The number of respondents that found a tick (n) and the total in each frequency per recreational outdoor activity are included (N), followed by the percentage (%). The odds of finding a tick when doing an activity about once a summer, monthly or weekly compared to never doing the activity was calculated, the odds ratio (OR) and 95% confidence interval (95% CI) are included. A comparison between the Midwest and Northeast was made for participants that reported finding a tick versus those who did not, the p-value of the Chi-squared test (χ^2^) is included.

### Peridomestic risk factors

Of all houses with a yard, nearly all had a manicured lawn and outdoor seating, and approximately 25% had children’s play equipment (Figure 2A). Properties in Wisconsin had more birdfeeders (χ^2^ test, n=892, df=1, χ^2^=524.46, p<0.001) and log or brush piles (χ^2^ test, n=891, df=1, χ^2^=36.106, p<0.001), whereas fences were more common in the Northeast (χ^2^ test, n=890, df=1, χ^2^=42.368, p < 0.001) (Figure 2A). Self-reported deer sightings (never or rarely versus more frequent sightings) on properties with a yard were not significantly different between respondents from the Midwest and the Northeast (χ^2^ test, n=893, df=1, χ^2^=0.159, p=0.69). Daily sightings happened at 16.3% (100 of 615 Midwest participants) and 14.0% of homes (39 of 278 Northeast participants), while deer were never or rarely observed at 47.5% (Midwest) and 48.9% (Northeast) of homes. Deer proof fences (χ^2^ test, n=892, df=1, χ^2^=30.593, p<0.001) and deer resistant plants (χ^2^ test, n=888, df=1, χ^2^=17.867, p<0.001) were more commonly used among participants from the Northeast. By contrast, baiting to attract deer to yards was more common among participants from the Midwest (χ^2^ test, n=890, df=1, χ^2^=8.140, p=0.004) although still infrequent (5.7% of 615 versus 1.5% of 275, Figure 2B).

**Figure 2:**
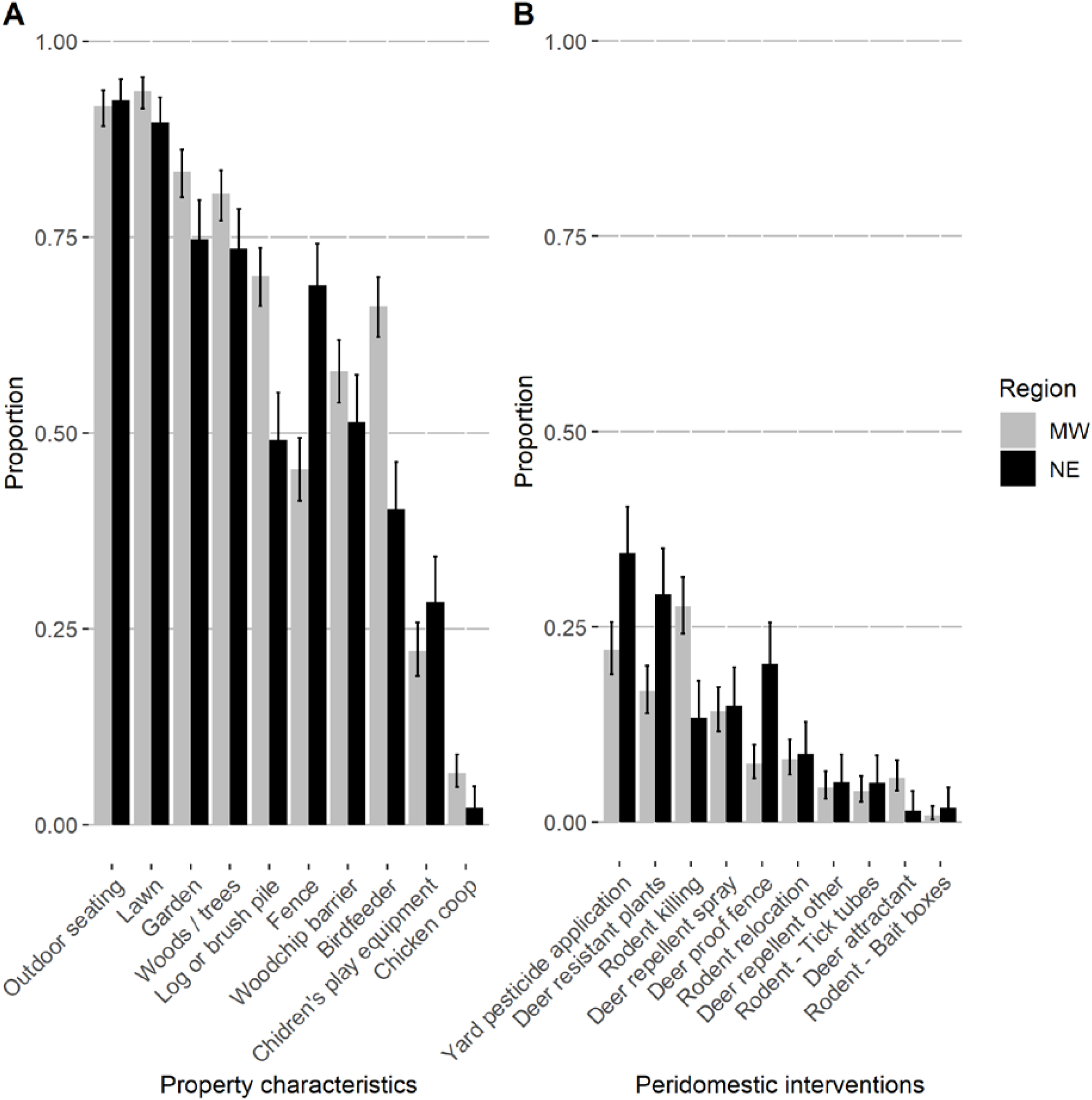
Peridomestic risk factors for tick exposure were more prevalent in the Midwest and peridomestic tick interventions were more common in the Northeast. A) The proportion of homes (excluding apartments or condominiums) from participants from the Midwest (grey) and Northeast (black) with each property characteristic. B) Peridomestic interventions employed in the Midwest and Northeast targeting deer, rodents and the environment. Error bars represent 95% confidence intervals.

Rodent-targeted interventions to control ticks were rarely used; less than 5% of participants used tick tubes or bait boxes (Figure 2B). Killing rodents (i.e. chipmunks and mice) on the property was much more common among respondents from the Midwest (χ^2^ test, n=889, df=1, χ^2^=21.772, p<0.001) (Figure 2B); of the Midwesterners who killed nuisance rodents, 18.9% reported doing this at least weekly (n=32). Use of pesticides targeting ticks, mosquitoes, or other insects on the property was more common among Northeasterners (χ^2^ test, n=895, df=1, χ^2^=15.205, p<0.001) (Figure 2B). However, among those who applied pesticides, the frequency of application did not differ between the two study areas (χ^2^ test, n=228, df=2, χ^2^=2.185, p=0.3). Seasonal (45.2%) and monthly (48.2%) applications were most commonly reported and weekly application was rare (6.6%).

Participation in peridomestic activities was common among participants residing on properties with a yard; 49.9% of 888 participants reported mowing the lawn weekly and 61.7% of 888 gardened at least weekly. The frequency of mowing the lawn and gardening was higher among respondents from the Midwest than the Northeast (Table 2). Participants who reported weekly gardening or lawn mowing in spring and summer were at least twice as likely to report that they had found a tick on themselves the previous fall and winter compared to participants who never gardened or mowed the lawn (Table 3).

### Modeling personal protective behaviors

For each of the recreational outdoor activities, the likelihood of using the four most common personal protective behaviors (check themselves for ticks, adjust their clothing, use repellent, or shower and bathe to remove ticks) increased with the reported frequency of the activity during the spring and summer, after adjusting for the region, gender, age category, and previous Lyme disease diagnosis (Figure 3A). There was strong evidence for the relationship between personal protective behaviors and going to the beach, birding, camping, fishing, and hiking (Figure 3A, p<0.001, Supp. Table 1) and moderate evidence for the relationship with hunting (Figure 3A, p-value range: 0.01 – 0.02, Supp. Table 1). Participants who reported doing more frequent peridomestic outdoor activities were more likely to use any of the four common prevention strategies if they gardened (Figure 3B, p-value range: 0.015-0.026, Supp. Table 2), after accounting for region, previous Lyme disease diagnosis, age, gender, frequent participation in recreational outdoor activities (outdoor index), and the use of deer, rodent, and/or environmental control measures. However, mowing the lawn was not associated with use of personal protective behaviors (Figure 3B, p-value range: 0.74-0.98, Supp. Table 2). In this peridomestic scenario, participants who reported any type of deer control, also reported using three of the four most common personal protective behaviors (check for ticks, adjust clothing, and shower and bathe to remove ticks) more frequently (Supp. Table 2). The use of rodent and environmental control measures was not associated with use of personal protective behaviors.

**Figure 3:**
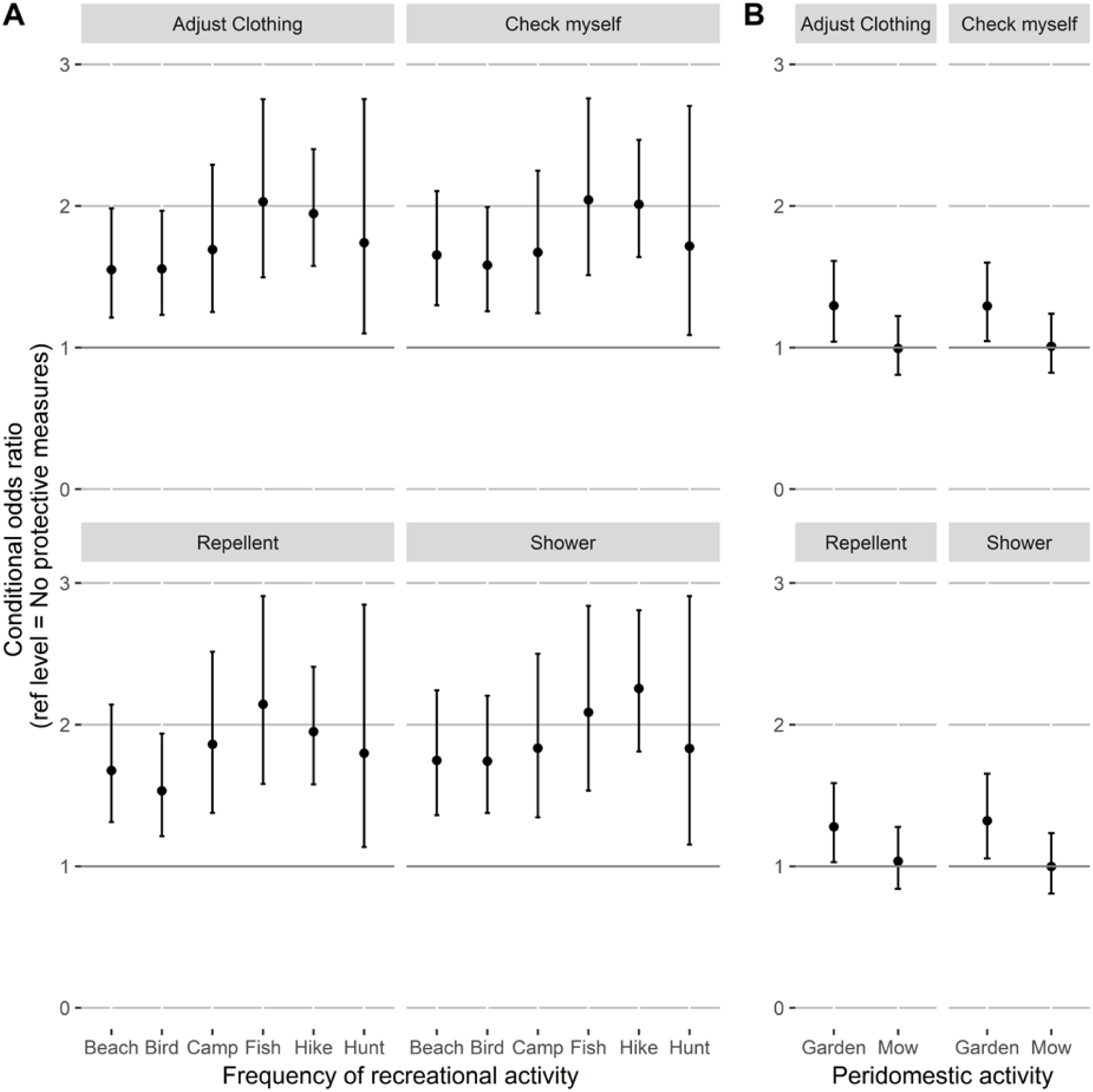
More frequent recreational and peri-domestic activities, except mowing the lawn, were associated with the use of personal protective measures. The conditional odds ratios represent the conditional estimate for increased outdoor activity and the likelihood of use of a preventative measure. A) Estimates for recreational outdoor activities, after accounting for age category, gender, the interaction between activity frequency and region, and previous Lyme diagnosis. B) Estimates for peridomestic activities. In addition to the previously mentioned model parameters, the model also accounted for peridomestic interventions for deer, rodents and insecticide treatment, and if participants did frequent recreational outdoor activities. Error bars represent 95% confidence interval.

The likelihood of use of personal protective behaviors among respondents from the Northeast was about half compared to Midwesterners for those who went birding (cOR range: 0.51-0.57, p-range: 0.003-0.01, Supp. Table 1), visited the beach (cOR range: 0.43-0.51, p-range: <0.001-0.007, Supp. Table 1), went hunting (cOR range: 0.49-0.56, p-range: 0.002-0.012, Supp. Table 1), or went camping (cOR range: 0.54-0.62, p-range: 0.01-0.05, Supp. Table 1) after accounting for region, previous Lyme disease diagnosis, age and gender. That is, people with the same risk of exposure reported using fewer personal protective behaviors in the Northeast except for those that fished, hiked, gardened or mowed the lawn. In the outdoor activity models, there is marginal evidence of increased reporting of personal protective behaviors with a previous Lyme disease diagnosis after we accounted for the aforementioned covariates (cOR 1.296-2.475, p-range: 0.054-0.492, Supp. Table 1 and 2).

## Discussion

Behavioral and residential risk factors for exposure to ticks and tick-borne diseases were different for study participants from the Northeast and Midwest. Midwesterners participated at a greater rate and more frequently in recreational outdoor activities (81.9% vs 63.1%). Similarly, Midwesterners were more likely to engage in the peridomestic activities of weekly lawn mowing (60.5% vs 26.4%) and gardening (65.8% vs 52.7%) compared to Northeasterners. These observations suggest that exposure to ticks and tick-borne diseases could be higher for Midwestern study participants compared to Northeastern participants. Indeed, the frequency of tick encounters in the prior fall and winter was higher for Midwestern participants (37.3%) than Northeastern participants (22.0%). Midwesterners also utilized personal protective behaviors more frequently (Figure 1), perhaps as an adaptive behavior in response to higher risk of encountering a tick or other biting insects due to more frequent outdoor activities. In contrast, Northeasterners were more likely to use peridomestic interventions that could reduce tick and deer activity in their yards.

Overall, the use of personal protective behaviors was associated with more frequent participation in outdoor activities. Nonetheless, Northeasterners were less likely to use personal protective behaviors even when they were engaged in similar outdoor activities. This difference is opposite to the trend observed in the national HealthStyles survey, where participants from the Mid-Atlantic region, including New York, New Jersey and Pennsylvania, reported slightly higher use of personal prevention measures than respondents from East-North Central US, including Wisconsin, Michigan, Illinois, Indiana and Ohio (Hook et al., 2015). Although the two regions in the HealthStyles survey had similar reports of tick exposure (24% in Mid Atlantic to 22.8% in East-North Central), the East-North Central region included four states-Michigan, Illinois, Indiana and Ohio-where Lyme disease incidence rates are much lower and blacklegged ticks less common than in Wisconsin (Eisen et al., 2016). This lower risk of Lyme disease in most of the region may have resulted in the slightly reduced use of personal protective behaviors observed in the East North Central US survey responses.

Use of prevention measures generally increases if they are perceived as effective and not burdensome (Butler et al., 2016). The proportion of participants that reported checking themselves for ticks (74.3 and 86.7%) was high compared to reports from other high-incidence states, for example 30.7% in the Mid Atlantic (Hook et al., 2015) and 57-67% in Connecticut, (Connally et al., 2009; Niesobecki et al., 2019). The high percentage of checking for ticks in our study could be due to the profile of our study participants, who were more likely to be active outdoors and to have been diagnosed with tick-borne disease compared to the general public (Fernandez et al., 2019). Alternatively, this high percentage may have been an artifact of the structure of the survey question, because checking for ticks was the first option in the response list. While “checking for ticks” was reported more frequently than expected based on prior studies, the use of adjusting clothing (50 and 57.6%), wearing repellent (50.4 and 62.4%), or showering to remove ticks (38.8 and 52%) in our two study areas fell within the wide range of reported use percentages in high Lyme disease incidence states (Connally et al., 2009; Herrington, 2004; Niesobecki et al., 2019). Similar to Connally et al. (2009) and Niesobecki et al. (2019), the use of permethrin-treated clothing was least reported. However, there appears to be an increasing trend in the sales of permethrin-treated clothing (Online access Market Research Engine), which aligns with the increase in positive responses seen in surveys asking about the use of permethrin-treated clothing, from 0.7% in 2005-2007 (Connally et al., 2009), to 7% in 2015 (Niesobecki et al., 2019) and 14% in our study. Greater awareness and increased belief in the effectiveness together with increased availability of these products could explain this increase.

The observed differences in peridomestic risk factors and intervention strategies between the Northeastern and Midwestern participants might be due to differences in socio-ecological contexts and possibly influenced by differing urbanization levels between the regions compared. Southern New York and New Jersey are metropolitan areas whereas nearly two-third of Wisconsin counties (46 of 72) are nonmetropolitan (less than 50,000 inhabitants per urban cluster, (Ingram and Franco, 2014). Midwest participants represented a social group that is more active outdoors and also actively recruited wildlife like birds and deer into their yards. Birdfeeders, known to attract deer, squirrels, and other rodents, and log and brush piles, known to provide nesting space for rodents, were more common in the Midwest. These features might increase rodent activity on the property, which could explain why killing of rodents on properties was also more common in the Midwest than in the Northeast. In addition, while Midwest participants were more likely to provide forage for deer, deer proof fencing and deer resistant plants were more common in the Northeast. Despite the difference in the use of deer-targeted interventions, deer sightings on the properties were not significantly different between study regions.

Interestingly, the largest difference in outdoor activity frequency was related to mowing the lawn. Only 26.4% of Northeasterners with a yard did this weekly compared to 60.5% of Midwesterners. We posit that the difference in weekly lawn mowing participation is because lawn care companies do the work in the Northeast (unpublished data Fernandez & Diuk-Wasser). The use of a lawn care company could also explain the higher proportion of study participants who reported pesticide use on Northeastern properties compared to Midwestern properties. Taken together, the higher use of peridomestic interventions of Northeastern participants (i.e. reducing adult blacklegged tick hosts, deer, in the yard and using pesticides to kill ticks to reduce the environmental hazard) might suggest that they are more prone to invest in protecting their landscaping or have greater awareness of Lyme disease ecology compared to Midwestern participants. Further studies are needed to dissect these associations and their relationship to the socio-ecological contexts (e.g., risk perception, income).

Our study was a retrospective survey, with voluntary participation and mostly passive recruitment. This strategy resulted in a biased study population with participants who likely had an above average interest in preventing tick-borne diseases. In this study, we did not explicitly address differences in urbanization levels that could be driving the observed differences in behavioral and residential risk factors. This uneven representation of the population living in more urban areas in the Northeast *versus* the Midwest, warrants further exploration as more data becomes available from exurban and rural areas in the Northeast. In addition, any retrospective survey is subject to recall bias (Groves et al., 2011); increasing the frequency of reporting can reduce this bias with information recalled for a shorter period of time (Clarke et al., 2008). In The Tick App, participants can share their daily outdoor activities, personal prevention methods used, and tick encounters in less than a minute using the Daily Log functionality. This feature, called Tick diary in 2018, was sparingly used by Tick App users (Fernandez et al. 2019), but we anticipate that these daily assessments can provide more accurate estimates of prevention method use in relation to outdoor activities when a more robust sample is acquired. Lastly, although we tried to avoid “check all that apply” question structures where participants are more likely to select the first options (Rasinski et al., 1994), the personal preventative behavior question was structured this way. Possible over reporting of the first option (i.e. checking for ticks) and under reporting of later options could have been reduced by a forced reply structure (yes/no) or randomizing the order of responses, but randomizing was not possible in our smartphone application. In the 2019 Tick App enrollment survey, the “check all that apply” structure was replaced with a yes/no response structure.

Human behavior converts enzootic hazard into Lyme disease risk. The results of this study illustrate how personal prevention measures were more likely to be employed by participants in different socio-ecological contexts. To be able to include the human behavioral component into predictions of Lyme disease risk, researchers will need to determine how specific behaviors (activities and prevention strategies) relate to tick encounters and disease risk, across regions. The use of a smartphone application to deliver standardized surveys proved to be a cost-efficient tool to collect data regarding risk factors for Lyme disease and offers the opportunity of expanding the geographic scope of these studies. Ultimately, this information will be valuable to adapt risk reduction interventions and for generating tailored public health messages for different populations across regions in the United States.

## Data Availability

Summarized data is available upon request.

## Acknowledgements

We thank all participants for taking time to share their experiences and we thank everybody who promoted The Tick App. We extend our gratitude to our colleagues, beta-testers, UW-Madison Survey Center, Becky Eisen and Alison Hinckley for constructive feedback on the survey. Funding: This work was Supp.orted by Center for Disease Control and Prevention [Cooperative Agreements #U01 CK000505 and #U01 CK000509-01]. Its contents are solely the responsibility of the authors and do not necessarily represent the official views of CDC or the Department of Health and Human Services.

